# Balloon Angioplasty as the First-Choice Treatment for Intracranial Atherosclerosis-Related Emergent Large Vessel Occlusion Involving the Microcatheter “First-Pass Effect”

**DOI:** 10.1101/2023.01.30.23285210

**Authors:** Liang Zhang, Xiongjun He, Kaifeng Li, Li Ling, Min Peng, Li’an Huang, Yajie Liu

## Abstract

**Background:** It is unknown whether balloon angioplasty can be a first-choice treatment for intracranial atherosclerosis-related emergent large vessel occlusion (ICAS-ELVO) with small clot burden. The microcatheter “first-pass effect” is a valid predictor of ICAS-ELVO with small clot.

**Objective:** To determine balloon angioplasty’s efficacy as first-choice treatment for ICAS-ELVO involving the microcatheter “first-pass effect” during endovascular treatment (EVT).

**Methods:** This continuous retrospective analysis assessed ICAS-ELVO patients presenting with the microcatheter “first-pass effect” during EVT. Patients were divided into two first-choice treatment-based groups: preferred balloon angioplasty (PBA) and preferred mechanical thrombectomy (PMT). Efficacy and safety outcomes were compared between groups.

**Results:** Seventy-six patients with ICAS-ELVO involving the microcatheter “first-pass effect” during EVT were enrolled. Compared with patients in PMT group, patients in PBA group were associated with (i) a higher rate of first-pass recanalization (54.0% vs. 28.9%, p=.010) and complete reperfusion (expanded thrombolysis in cerebral ischemia≥2c; 76.0% vs. 53.8%, p=.049), (ii) a shorter puncture-to-recanalization time (49.5 min vs. 56.0 min, p<.001), (iii) less operation costs (48,499.5¥ vs. 99,086.0¥, p<.001),and (iv) more excellent functional outcomes (modified Rankin scale:0-1; 44.0% vs. 19.2%, p=.032) at 90 days. No significant differences in symptomatic intracranial hemorrhage (12.0% vs. 15.4%, p>.999) and mortality (10.0% vs. 7.7%, p>.999) were noted. Logistic regression analysis revealed that first-choice treatment was an independent predictor of 90-day excellent functional outcomes (adjusted odds ratio [aOR] =0.10, 95% CI: 0.02–0.66, p=.017).

**Conclusion:** Balloon angioplasty, as the first-choice treatment, potentially improves 90-day functional outcomes for ICAS-ELVO patients with microcatheter “first-pass effect” during EVT.

**What is already known on this topic:** Compared with large vessel occlusion caused by embolization, mechanical thrombectomy has lower recanalization rate, longer procedure time, and poorer prognosis for patients with intracranial atherosclerosis-related emergent large vessel occlusion(ICAS-ELVO).

**What this study adds:** This study revealed that balloon angioplasty, as the first-choice treatment, potentially improves 90-day outcomes, shortens procedure time, and reduces operation costs for patients with ICAS-ELVO involving the microcatheter “first-pass effect” during endovascular treatment.

**How this study might affect research, practice or policy:** We believe that our study makes a significant contribution to the literature because its findings suggest that rapid and accurate methods of diagnosing the etiology and clot burden of ELVO as well as the development of an individualized EVT strategy based on etiology and clot burden need to be established.

## INTRODUCTION

Mechanical thrombectomy (MT) has been the gold-standard of reperfusion therapy for patients with emergent large vessel occlusion (ELVO); however, these studies’ participants predominantly originated from Western populations, and the primary etiology of vascular occlusion was embolism ^1-5^. When treating intracranial atherosclerosis-related emergent large vessel occlusion (ICAS-ELVO), MT results in a lower recanalization rate, longer procedure times, and poorer outcomes than when treating embolism-induced vessel occlusion, and a significant number of patients require rescue therapy ^6-9^. Patients with ICAS-ELVO encounter unique technical challenges during endovascular recanalization therapy.

ICAS-ELVO is related to in-situ atherothrombotic occlusion and usually has a smaller clot burden ^10^, for which MT is often inefficient. Moreover, additional and repeated MT tends to damage the vascular endothelium and increases the risk of reocclusion. Balloon angioplasty has proven safe in ICAS-ELVO rescue therapy following MT, and we speculate that it may be a better first-choice treatment than MT for ICAS-ELVO in patients with a small clot burden.

Though certain studies have demonstrated the feasibility of direct angioplasty for patients with ICAS-ELVO ^11-14^, these studies did not define the clot burden of the included patients. The microcatheter “first-pass effect” is considered a valid predictor of ICAS-ELVO^15^, and it often suggests an occluded vessel with a small clot burden. Thus, we conducted this retrospective study to determine whether balloon angioplasty could be the first-choice treatment for ICAS-ELVO in patients with small clot burden, which was defined through the microcatheter “first-pass effect” that presented during endovascular treatment (EVT).

## METHODS

### Patients

We reviewed the information of the consecutively collected ELVO patients who underwent EVT between October 2018 and December 2021 in Shenzhen Hospital of Southern Medical University. We enrolled patients who:(i) were aged >18 years, (ii) presented with isolated intracranial internal carotid artery (ICA), M1 of middle cerebral artery, intracranial vertebral artery or basilar artery occlusion, and (iii) the microcatheter “first-pass effect” was evaluated before EVT. Patients were divided into two first-choice treatment-based groups: preferred balloon angioplasty (PBA) and preferred mechanical thrombectomy (PMT). A flowchart of the selection process is shown in **Figure 1**.

**Figure 1.**
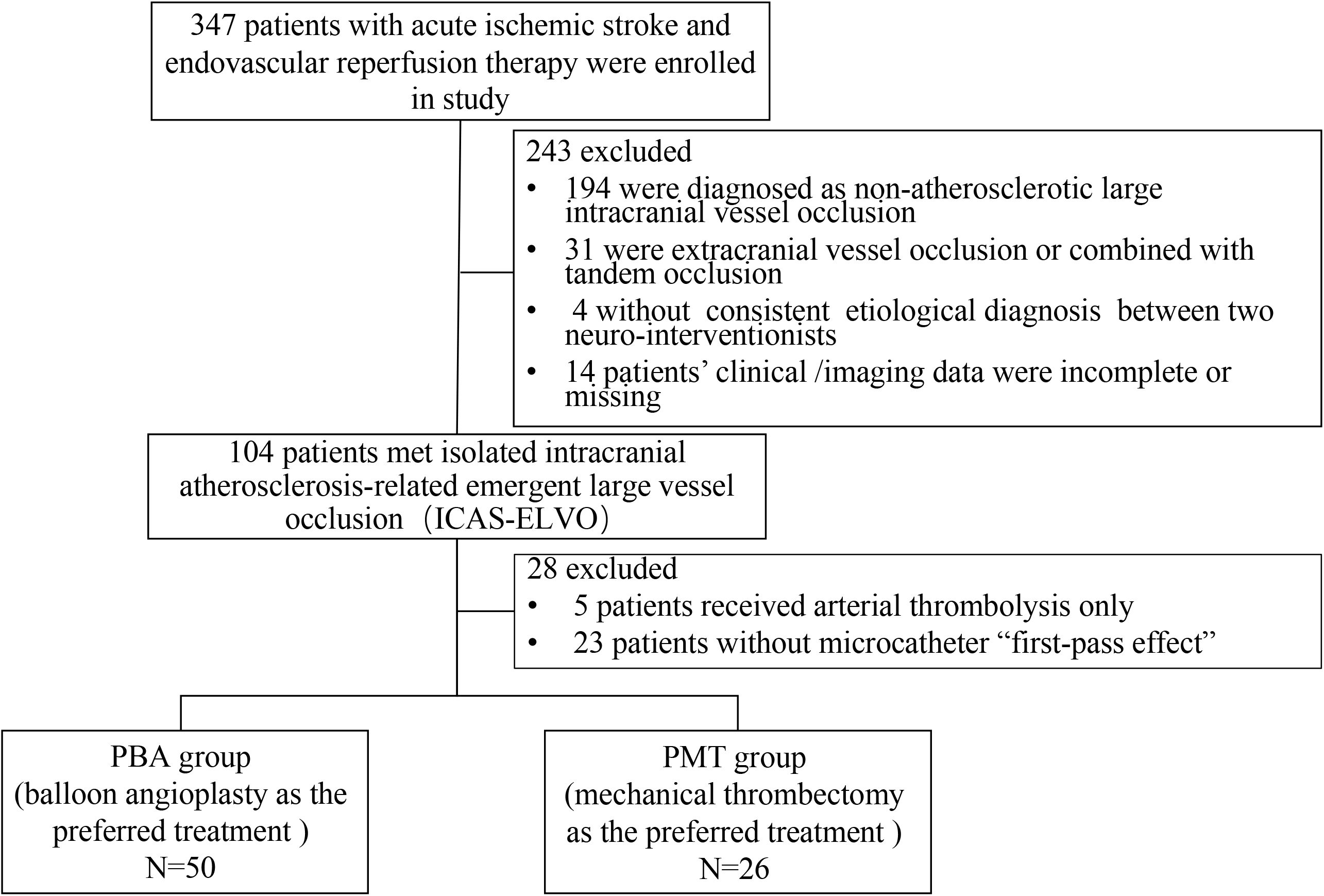
Flow chart of patient screening in this study

The etiologies of vessel occlusion and the microcatheter “first-pass effect” were assessed by two neurointerventionalists using clinical and imaging data. The definition of the microcatheter “first-pass effect” has been described in a previous study ^15^, and microcatheters used in this study mainly were the XT-27^®^ (Stryker Inc, Kalamazoo, MI, USA) and Rebar-27^®^ (Medtronic Inc, Minneapolis, MN, USA). Illustrative cases of the microcatheter “first-pass effect” are shown in **Figure 2**. The basis for determining ICAS-ELVO included: (i) clear evidence of ICAS as the culprit occluded vessel prior to this ischemic event; (ii) procedural evidence of ICAS after the first pass of the microcatheter or balloon dilation/stent deployment; and (iii) significant fixed focal stenosis (stenotic degree ≥ 50%) at the occlusion site. To reduce bias, both neurointerventionists were tested in 10 selected cases of ELVO whose etiology was identified as ICAS, and the diagnostic accuracy was 100%. This study was approved by our local medical ethics committee.

**Figure 2.**
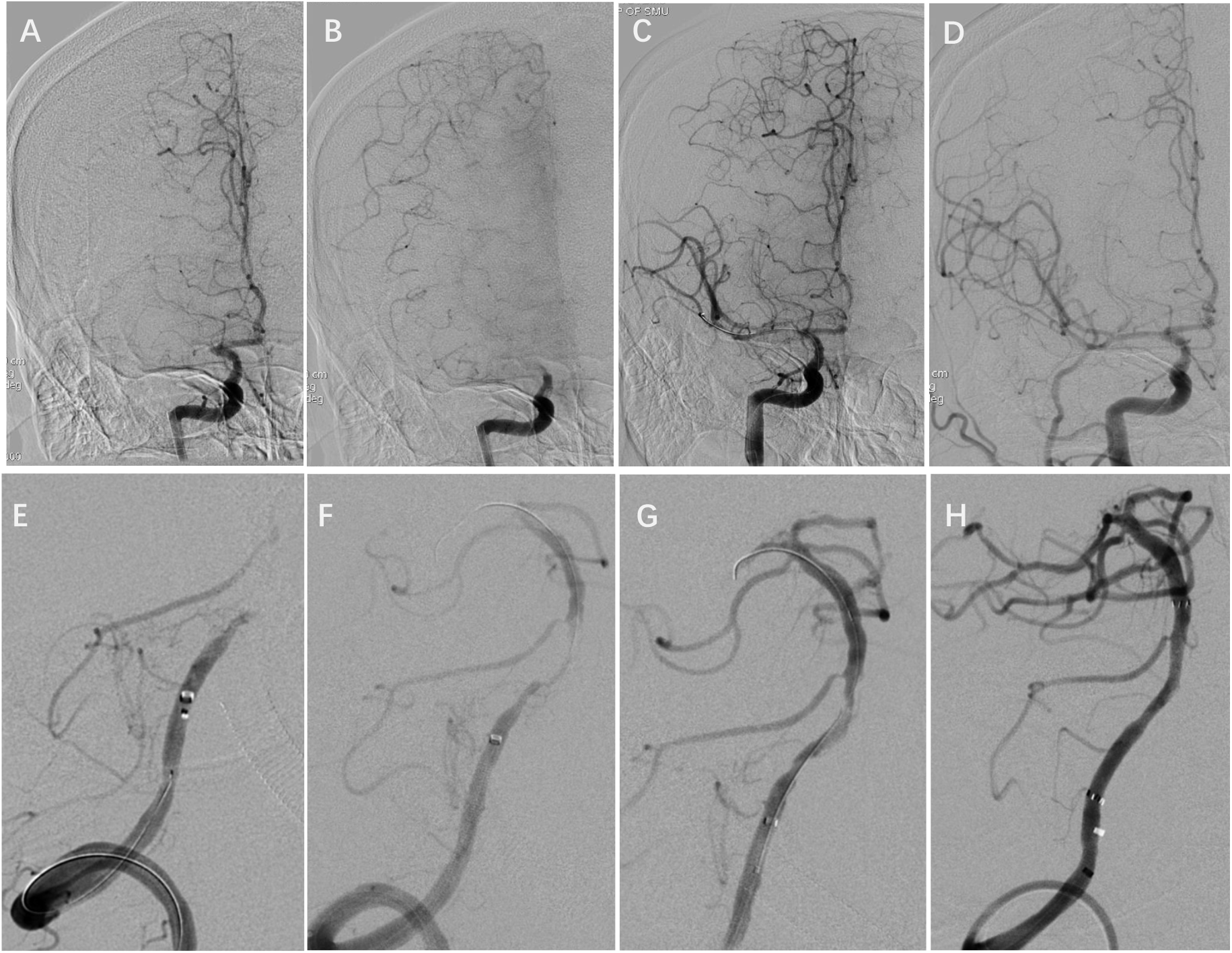
Illustrative cases of the microcatheter “first-pass effect.” (1): A 51-year-old male patient with hypertension and diabetes for 5 years presented with an acute onset of left body weakness (iNIHSS score: 9) for 11 h. Angiography revealed occlusion at the M1 segment of the right MCA and “taper sign” (**A**); Angiography exhibited adequate collateral, and retro-flow reconstituted the distal portion of M1 (**B**); An XT-27 microcatheter and a microwire were navigated through the occlusion area to the distal patent artery, and the microcatheter was subsequently withdrawn to the occlusion’s proximal side. Angiography demonstrated blood flow through the vessel at the occlusion site, which was defined as the microcatheter “first-pass effect” **(C)**; Direct balloon angioplasty (gateway 2.0 × 9.0 mm) achieved an mTICI score of 3 **(D)**. (2): A 65-year-old male patient with hypertension for 11 years presented with unconsciousness (iNIHSS score: 15) for 4 h. Angiography demonstrated that the V5 segment of the right dominant vertebral artery was occluded **(E)**; Angiography revealed the microcatheter “first-pass effect” **(F)**; Direct balloon angioplasty (Sino 2.25 × 15.0 mm) achieved an mTICI score of 2b with focal dissection **(G)**; An enterprise stent (4 × 39 mm) was placed at the stenotic site, and angiography revealed an mTICI score of 3 **(H)**.

### Endovascular Treatment

Our center is a national senior stroke center and has extensive experience in EVT. Patients with intravenous thrombolytic indications were administered t-PA intravenous thrombolytic therapy, and EVT was performed in patients with intravascular indications after they had provided written informed consent. Decisions regarding recanalization strategies were based on the neurointerventionalists’ discretion. When the first-choice treatment failed to recanalize or maintain vascular patency, rescue therapy was performed at the appropriate time. The choice of balloon size depended on the diameter measured at the proximal segment of the occluded vessel, and the selected balloon size was generally smaller than the vessel’s actual measured diameter. The balloon was gradually pressurized using a pressure pump, and the maximum pressure did not exceed the balloon’s pressure. Patients with inadequate preparation for oral antiplatelet agents received tirofiban before balloon angioplasty and/or stenting. All operations were performed by neurointerventionalists who had > 5 years’ EVT experience.

### Clinical Characteristics and Outcome Assessment

The demographic, clinical, and imaging data of patients were collected from database of the hospital. A head computed tomography (CT) scan was performed within 24 h after surgery to determine the presence of intracranial hemorrhage. Head CT scans were reviewed at any time in patients with early neurological deterioration.

The primary efficacy outcome was a postoperative 90-day excellent functional outcome (modified Rankin scale [mRS: 0–1]), which was evaluated at 90 days follow-up reexamination by neurologists who were not involved in the procedure, and telephone-based follow-up was conducted if the patient could not return to the hospital. Secondary efficacy outcomes included 90-day functional independence (mRS 0–2), first-pass recanalization (FPR) rate, successful recanalization rate and complete reperfusion rate. FPR is defined as a single thrombectomy, or balloon angioplasty performed to achieve successful recanalization without the use of rescue therapy. Successful recanalization was defined as grade 2b–3 expanded thrombolysis in cerebral ischemia (eTICI), and complete reperfusion was defined as grade 2c–3 eTICI ^16 17^. The safety endpoints were symptomatic intracranial hemorrhage and mortality within 24 h and 90 days after surgery, respectively. Intracranial hemorrhage was assessed using CT, and symptomatic intracranial hemorrhage was defined as neurological deterioration and a ≥ 4-point increase in the National Institutes of Health Stroke Scale score on admission (i-NIHSS) ^18^.

### Statistical Analysis

SPSS 20.0 software (IBM Inc, Armonk, NY, USA) was used for statistical analysis. Categorical variables are presented as frequencies (%) and non-normally distributed continuous variables as medians (interquartile ranges [IQRs]). Between-group differences in clinical and procedural characteristics were compared using the χ^2^-test, Student’s t-test, Fisher’s exact test, or Mann-Whitney U depending on the type of variables analyzed. To evaluate independent predictors of excellent functional outcome at 90 days, we performed a logistic regression model with the following terms in the model: first-choice treatment, i-NIHSS, age, sex, and any other baseline characteristic with a probability value ≤ 0.10 in univariate analyses. All tests were two-tailed, and statistical significance was set at p≤0.05.

## RESULTS

### Baseline Characteristics and Procedural Details

A total of 76 patients were included in the study. Regarding the preferred treatment, 50 patients underwent balloon angioplasty (PBA group), and 26 underwent MT (PMT group). In the PBA group, 46.0% (23/50) patients underwent stenting and/or thrombectomy (13 patients underwent stenting only, five underwent thrombectomy only, and five underwent both stenting and thrombectomy). In the PMT group, 76.9% (20/26) patients underwent balloon angioplasty and/or stenting (four patients underwent balloon angioplasty only, one underwent stenting only, and 15 underwent both balloon angioplasty and stenting).

Baseline characteristics and procedural details are summarized in Table 1. No significant differences were noted in age, sex, hypertension, diabetes, hyperlipidemia, coronary artery disease, smoking, history of stroke, i-NIHSS, intravenous thrombolysis, glycoprotein IIb/IIIa inhibitor use, general anesthesia, location of vessel occlusion, and time from puncture to first recanalization between the two groups. However, the time from puncture to recanalization (49.5 min vs. 56.0 min, p<.001), rescue therapy rate (46.0% vs. 76.9%, p=.010), and operation costs (48,499.5 ¥ vs. 99,086.0 ¥, p<.001) were significantly lower in the PBA group than in the PMT group.

**Table 1.** Comparison of baseline characteristics and procedural details between the two groups

|  | Overall<br>(n=76) | PBA group<br>(n=50) | PMT group<br>(n=26) | P value |
| --- | --- | --- | --- | --- |
| Age (y), mean (±SD) | 64.9±10.7 | 64.6±10.7 | 65.7±10.8 | 0.660 |
| Male, n (%) | 58 (76.3) | 39 (78.0) | 19 (73.1) | 0.632 |
| Hypertension, n (%) | 59 (77.6) | 41 (82.0) | 18 (69.2) | 0.205 |
| Diabetes mellitus, n (%) | 33 (43.4) | 20 (40.0) | 13 (50.0) | 0.404 |
| Hyperlipidemia, n (%) | 30 (39.5) | 18 (36.0) | 12 (46.2) | 0.390 |
| Coronary artery disease, n (%) | 9 (11.8) | 6 (12.0) | 3 (11.5) | 0.633 |
| Smoking, n (%) | 20 (26.3) | 16 (32.0) | 4 (15.4) | 0.171 |
| History of stroke, n (%) | 21 (27.6) | 15 (30.0) | 6 (23.1) | 0.522 |
| Location of occlusion site, n (%) |  |  |  | 0.532 |
| Anterior circulation | 49 (64.5) | 31 (62.0) | 18 (69.2) |  |
| Posterior circulation | 27 (35.5) | 19 (38.0) | 8 (30.8) |  |
| i-NIHSS, median (IQR) | 10 (8-13) | 10 (8-13) | 10 (8-14) | 0.930 |
| IV-tPA, n (%) | 47 (61.8) | 30 (60.0) | 17 (65.4) | 0.647 |
| Glycoprotein IIb/IIIa inhibitor use, n (%) | 54 (71.1) | 35 (70.0) | 19 (73.1) | 0.779 |
| General anesthesia, n (%) | 38 (50.0) | 26 (52.0) | 12 (46.2) | 0.629 |
| Means of recanalization, n (%) |  |  |  |  |
| Stent retrieval | 30 (39.5) | 7 (14.0) | 23 (88.5) | <0.001** |
| Aspiration | 7 (9.2) | 3 (6.0) | 4 (15.4) | 0.222 |
| Balloon dilation | 69 (90.8) | 50 (100.0) | 19 (73.1) | <0.001** |
| Stenting | 34 (44.7) | 18 (36.0) | 16 (61.5) | 0.034** |
| Puncture-to-first recanalization time (min), median (IQR) | 35.0 (29-42.5) | 34.5 (27.0-41.0) | 38.5 (33.0-45.0) | 0.074 |
| Puncture-to-recanalization time (min), median (IQR) | 56.0 (34.5-85.5) | 49.5 (31.0-70.0) | 89.0 (70.0-110.0) | <0.001** |
| Rescue therapy, n (%) | 43.0 (56.6) | 23 (46.0) | 20 (76.9) | 0.010** |
| Costs (¥), median (IQR) | 70,069.5<br>(44,593.5-95,210.0) | 48,449.5<br>(42,749.0-77,478.0) | 99,086.0<br>(67,639.0-113,486.0) | <0.001** |

SD, standard deviation; i-NIHSS, National Institute of Health Stroke Scale score at admission;
IQR, interquartile range; IV-tPA, intravenous tissue-type plasminogen activator; \*\* $p \leq 0.05$ .

### Recanalization and Clinical Outcome

Safety and efficacy endpoint comparisons between the two groups are shown in Table 2. Overall, 98.7% (75/76) patients had grade 2b–3 eTICI, and one patient in the PMT group failed to recanalize because of iatrogenic dissection. 65.8% (50/76) patients achieved 90-day functional independence (mRS≤2), and 35.5% (27/76) patients achieved 90-day excellent functional outcome (mRS≤1). The comparison of 90-day mRS score changes between the groups is shown in **Supplementary Figure 1**. No significant differences were observed in the successful revascularization rate (100% vs. 96.2%, p=.342), 90-day functional independence (72.0% vs. 53.8%, p=.551), symptomatic intracranial hemorrhage rate (12.0% vs. 15.4%, p>.999), and mortality rate (10.0% vs. 7.7%, p>.999) between the two groups. However, the rates of FPR (54.0% vs. 28.9%, p=.010), complete reperfusion (76.0% vs. 53.8%, p=.049), and 90-day excellent functional outcomes (44.0% vs. 19.2%, p=.032) were significantly higher in the PBA group than in the PMT group.

**Table 2.**
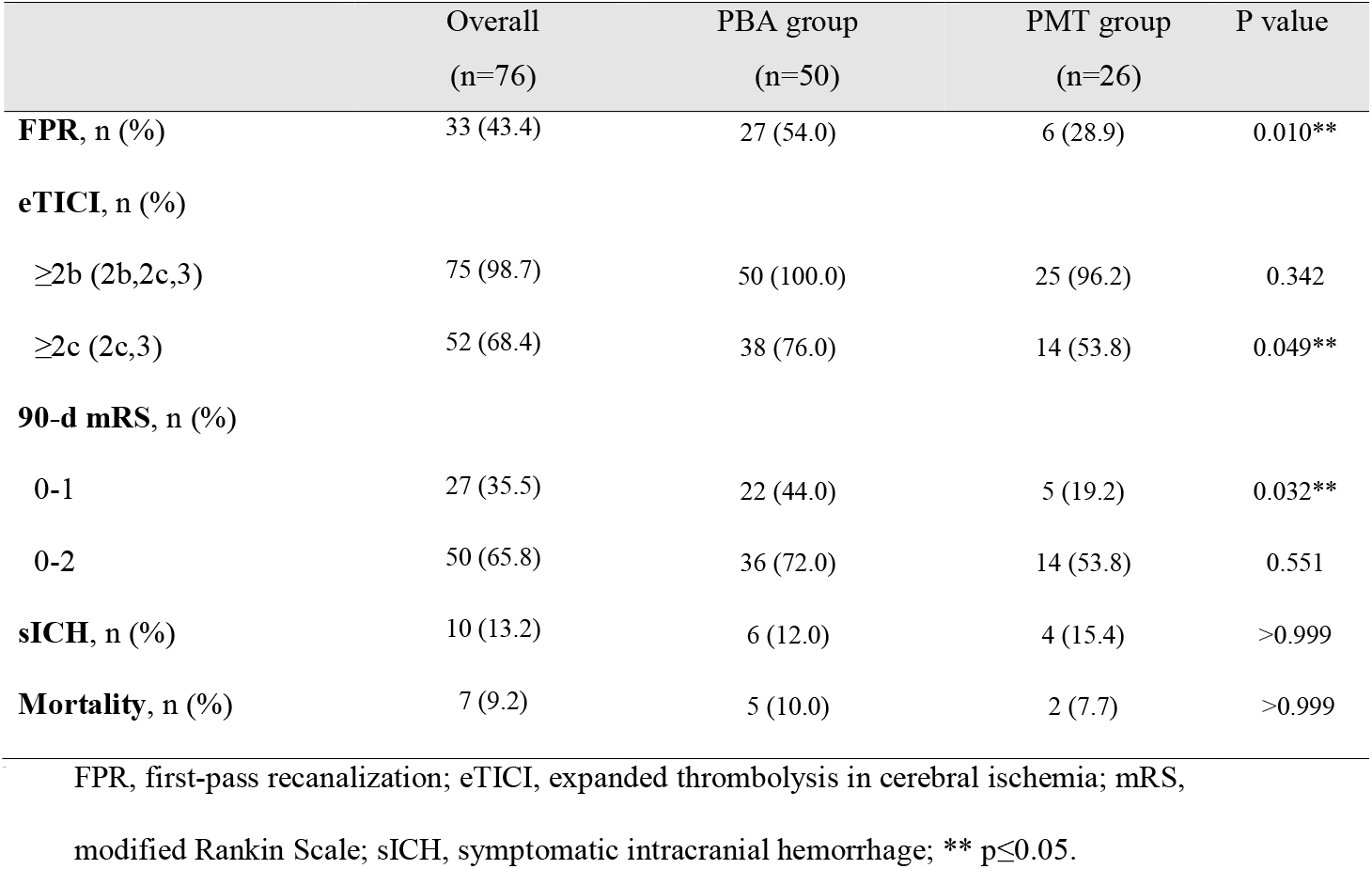
Efficacy and safety endpoint comparisons between the two groups

|  | Overall<br>(n=76) | PBA group<br>(n=50) | PMT group<br>(n=26) | P value |
| --- | --- | --- | --- | --- |
| **FPR, n (%)** | 33 (43.4) | 27 (54.0) | 6 (28.9) | 0.010** |
| **eTICI, n (%)** |  |  |  |  |
| ≥2b (2b,2c,3) | 75 (98.7) | 50 (100.0) | 25 (96.2) | 0.342 |
| ≥2c (2c,3) | 52 (68.4) | 38 (76.0) | 14 (53.8) | 0.049** |
| **90-d mRS, n (%)** |  |  |  |  |
| 0-1 | 27 (35.5) | 22 (44.0) | 5 (19.2) | 0.032** |
| 0-2 | 50 (65.8) | 36 (72.0) | 14 (53.8) | 0.551 |
| **sICH, n (%)** | 10 (13.2) | 6 (12.0) | 4 (15.4) | >0.999 |
| **Mortality, n (%)** | 7 (9.2) | 5 (10.0) | 2 (7.7) | >0.999 |

FPR, first-pass recanalization; eTICI, expanded thrombolysis in cerebral ischemia; mRS, modified Rankin Scale; sICH, symptomatic intracranial hemorrhage; \*\* p≤0.05.

Multivariate logistic regression analysis revealed that the i-NIHSS and first-choice treatment (PMT vs. PBA) were independent factors for 90-day excellent functional outcomes (adjusted odds ratio [aOR]=.53, 95% confidence interval [CI]: .36–.76, p<0.001 and aOR=0.10, 95% CI: 0.02–0.66, p=.017, respectively). The results of the univariate and multivariate logistic regression analyses are shown in Supplementary Table 1.

**Supplementary Table 1.**
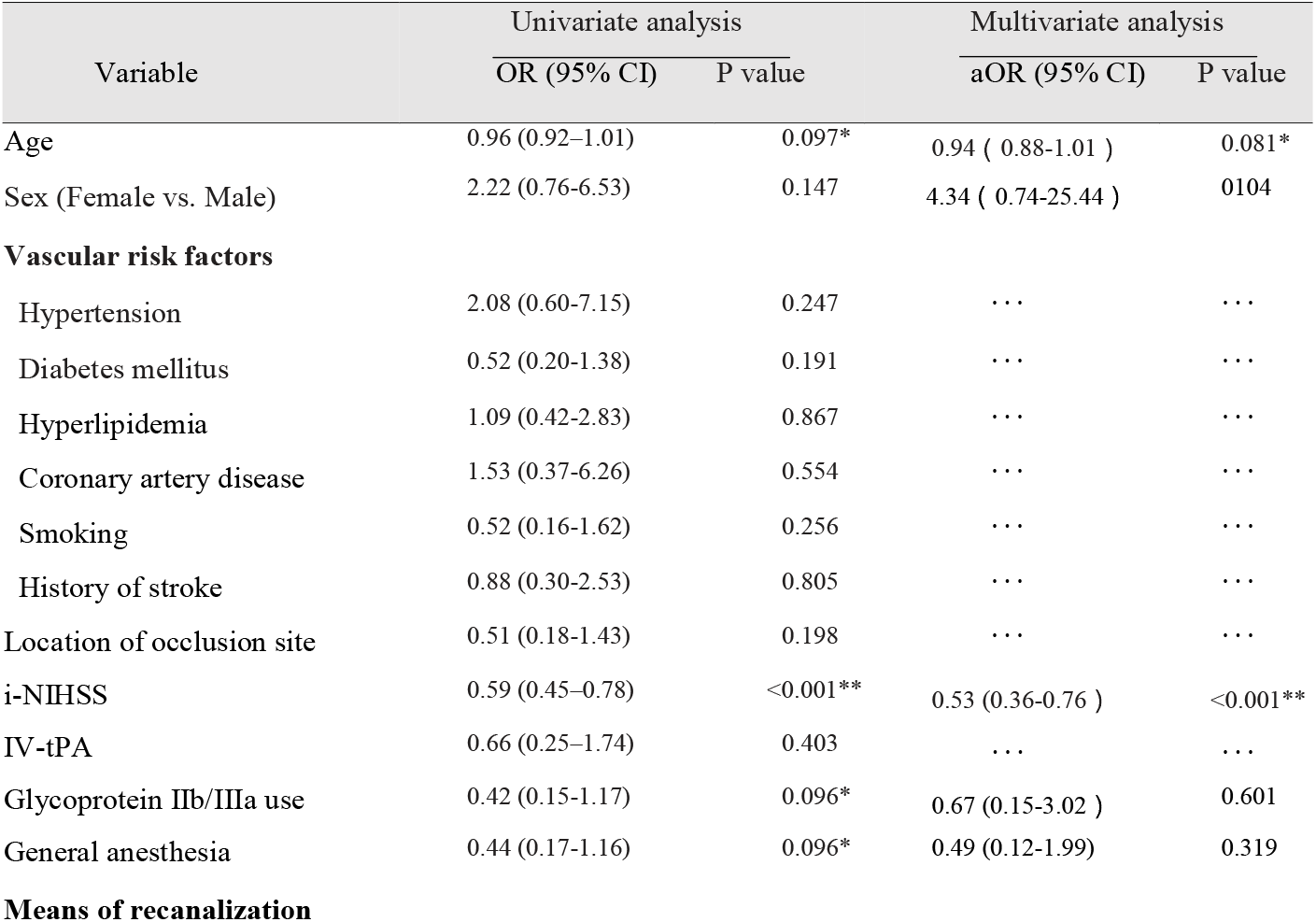
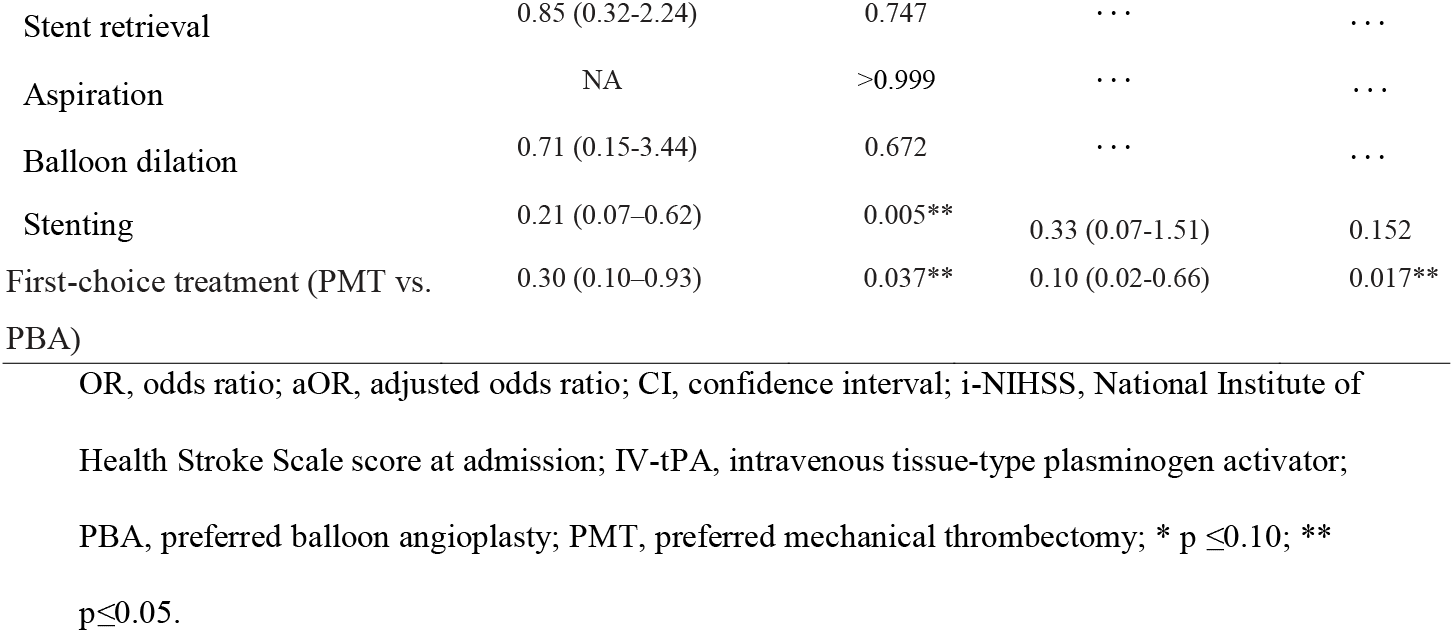
Logistic regression analysis for predicting 90-day excellent functional outcome (mRS≤1)

## DISCUSSION

For patients with ICAS-ELVO involving the microcatheter “first-pass effect” during EVT, two preferred treatments—MT and balloon angioplasty—were used for recanalization in this study. Our results indicate that the two treatments were comparable in terms of safety. However, they also show that balloon angioplasty, as the first-choice treatment, potentially increases the FPR and complete reperfusion rates, shortens the puncture-to-recanalization time, reduces the rescue therapy rate, and improves patients’ 90-day outcomes.

MT is often accompanied by a lower recanalization rate, longer operation time, and a higher risk of vascular reocclusion in patients with ICAS-ELVO, a phenomenon that has always challenged neurointerventionalists. Owing to the special pathophysiological characteristics of ICAS-ELVO, researchers are attempting to develop a targeted EVT strategy based on etiological diagnosis.

Certain studies have reported the advantages of direct angioplasty as a primary treatment for ICAS-ELVO, including shortening the procedure time and reducing the number of procedure device attempts, with similar safety to that of MT ^13 14^. However, these studies did not define the clot burden of the included patients, thus potentially affecting the persuasiveness of the results. ICAS-ELVO is related to in situ thrombotic occlusion, which results from unstable plaque ruptures and typically has a smaller clot burden; however, it is not absolute. Studies have demonstrated that clot burdens are associated with patient prognosis after EVT ^19^, and balloon angioplasty, as the first-choice treatment for ICAS-ELVO, is often ineffective in cases of large clot burdens. Thus, considering the clot burden as one of the bases for developing an EVT strategy for patients with ICAS-ELVO is recommended. Although the clot burden score ^20 21^ has been used to evaluate clot burdens, its extensive and accurate evaluation in clinical practice remains challenging. The microcatheter “first-pass effect” is a valuable predictor of ICAS-ELVO based on the condition’s small clot size and relative ease of disintegration; when the microcatheter passes through the occlusive site, it pushes part of the fresh thrombus to the blood vessel’s distal end. Thus, the microcatheter “first-pass effect” may also be an effective indicator of a small clot burden and an important variable for planning the ICAS-ELVO EVT strategy. Rapid and complete reperfusion after thrombectomy is a predictor of favorable outcomes in ELVO stroke ^16 22^. However, in patients with ICAS-ELVO, vessels tend to reocclude after recanalization due to post-MT endothelial injury, and repeated MT and/or angioplasty must be performed, thus substantially delaying reperfusion. Moreover, despite achieving successful vessel recanalization after MT, a considerable number of patients fail to achieve complete reperfusion because of residual stenosis. In this study, we compared the rates of FPR, complete reperfusion (eTIC≥2c), and 90-day excellent functional outcomes (mRS≤1) between PBA and PMT patients with ICAS-ELVO involving the microcatheter “first-pass effect” during EVT. The results revealed that balloon angioplasty, as the first-choice treatment, potentially increases the FPR and complete reperfusion rates and shortens the puncture-to-recanalization time; moreover, the puncture-to-recanalization time and FPR rate were significantly correlated with 90-day excellent functional outcomes. Our study implies that, for patients with ICAS-ELVO, balloon angioplasty that entails selecting an appropriate balloon size and dilatation pressure for angioplasty may be less invasive to the vascular endothelium than MT, which is beneficial for maintaining vascular stability after the first recanalization.

EVT is recognized as an effective method for ELVO and has been increasingly promoted worldwide. However, cost is often an important consideration for families and patients when deciding on EVT, especially for self-funded patients with ICAS-ELVO. Given the complexity of MT for ICAS-ELVO, neurointerventionalists often tend to estimate relatively high surgical costs, thus further increasing the concern of the patient’s family. Previous studies have observed that direct balloon angioplasty can reduce the number of procedure device attempts in patients with ICAS-ELVO; however, its ability to alleviate surgical costs has not been discussed. Our results indicate that balloon angioplasty, as the first-choice treatment, potentially reduces the use of remedial measures, thus relieving the economic burden on patients with ICAS-ELVO. Considering that approximately one-third of ischemic strokes in Asia are caused by ICAS, ensuring the effectiveness of EVT while reducing the cost as much as possible will benefit a greater number of patients.

Emergency balloon angioplasty’s major cause for concern is its increased risk of reocclusion without adequate antiplatelet therapy or cerebral hemorrhage when antiplatelet therapy is administered within 24 h of intravenous thrombolysis. Tirofiban is a short-acting intravenous antiplatelet drug widely used in percutaneous coronary intervention, and studies have proven that the combined use of tirofiban during or before EVT is feasible in patients with ICAS-ELVO ^11 23-25^. Furthermore, tirofiban may be an alternative therapy for patients with post-MT ICAS-ELVO ^26^. In this study, we found that balloon angioplasty, as the first-choice treatment, neither increased tirofiban use nor amplified the risk of symptomatic intracranial hemorrhage. In contrast, tirofiban use was more aggressively required in the PMT group owing to endothelial injury caused by additional and repeat MT. Another cause for concern in balloon angioplasty is the “snowplowing” effect: branch occlusion following angioplasty may cause secondary ischemia in the injury site. However, because the territory supplied by the neighboring side branches is often already infarcted following the complete occlusion of the trunk vessel, the risk of secondary ischemic injury due to “snowplowing” is not as high as anticipated ^8^.

Planning a targeted EVT strategy based on etiological diagnosis is an attractive option; however, predicting the etiology of vascular occlusion promptly and accurately before EVT remains difficult. Currently, no widely accepted diagnostic criteria for distinguishing atherothrombotic ELVO from others and potential preprocedural methods for identifying signs of atherosclerosis are available. For example, patients repeatedly suffering from transient ischemic attack-related symptoms on the ipsilateral side before acute ischemic stroke, combined with risk factors associated with atherosclerosis, have exhibited lower NIHSS scores and favorable collateral circulation upon admission, which leads to the suspicion of ICAS-ELVO ^9^. Imaging predictors, such as a core infarction located in the watershed area, severe calcification at the site of occlusion, severe and/or multifocal narrowing on angiography, “tapered” occlusion ^27^, “truncal-type” occlusion ^28^, and the microcatheter “first-pass effect,” ^15^ all support ICAS as the most likely etiology. In the future, more diagnostic methods that enable the rapid and accurate diagnosis of vascular occlusion’s etiology and provide a basis for treatment decisions are warranted.

## Limitations

There were several limitations in this study. First, our findings should be cautiously interpreted, as the study is characterized by a small sample size, single center, and retrospective nature; therefore, further prospective, multicenter, large sample-sized studies are required. Second, although a small clot burden is the prerequisite for the existence of microcatheter “first-pass effect,” its specificity and sensitivity to predict small clot burden need to be further tested in more studies.

## CONCLUSION

Our study reveals that balloon angioplasty, as the first-choice treatment, is potentially beneficial in improving the rate of 90-day excellent functional outcomes, shortening the procedure time, and reducing surgical costs for patients with ICAS-ELVO involving the microcatheter “first-pass effect” during EVT. In the future, the rapid and accurate diagnosis of the etiology and clot burden of ELVO and development of an individualized EVT strategy based on etiology and clot burden will be the focus of our subsequent research.

## Data Availability

All data produced in the present study are available upon reasonable request to the authors

## Abbreviations

aOR: adjusted odds ratio
CI: confidence interval
CT: computed tomography
ELVO: emergent large vessel occlusion
eTICI: expanded thrombolysis in cerebral ischemia
EVT: endovascular treatment
FPR: first pass recanalization
ICAS: intracranial atherosclerosis
IQR: interquartile range
mRS: modified Rankin Scale
MT: mechanical thrombectomy
NIHSS: National Institutes of Health Stroke Scale
PBA: preferred balloon angioplasty
PMT: preferred mechanical thrombectomy
PRT: puncture to recanalization time
SD: standard deviation
t-PA: tissue-type plasminogen activator
sICH: symptomatic intracranial hemorrhage.

## Acknowledgments

We are grateful to the clinicians and staff at the Shenzhen Hospital of Southern Medical University who contributed to the medical intervention and data collection for this study. We would like to thank Editage (www.editage.cn) for English language editing and Charlesworth (www.cwauthors.com.cn) for statistical review.

## Author Contributions

L.Z. and X.H. analyzed and interpreted the data and drafted the manuscript. Y.L. conceived and designed the research. M.P. and K.L. evaluated the image. L.L. and L.H. made critical revisions of the manuscript. All authors read and approved the final manuscript.

## Ethical Statement

The studies involving human participants were reviewed and approved by the Clinical Research Ethics Committee of Shenzhen Hospital of Southern Medical University. The patients/participants provided written informed consent to participate in this study.

## Data Availability Statement

The datasets used and/or analyzed during the current study are available from the corresponding author on reasonable request.

## FIGURE LEGENDS

**Supplementary Figure 1.** Comparison of modified Rankin Scale (mRS) scores at 90 days between the two groups

