## Supplementary Table 1 for "Balloon Angioplasty as the First-Choice Treatment for Intracranial Atherosclerosis-Related Emergent Large Vessel Occlusion Involving the Microcatheter “First-Pass Effect”"

**Supplementary Table 1** Logistic regression analysis for predicting 90-day excellent functional outcome (mRS≤1)

| Variable | Univariate analysis  OR (95% CI) P value | | Multivariate analysis  aOR (95% CI) P value | |
| --- | --- | --- | --- | --- |
| Age | 0.96 (0.92–1.01) | 0.097* | 0.94（0.88-1.01） | 0.081* |
| Sex (Female vs. Male) | 2.22 (0.76-6.53) | 0.147 | 4.34（0.74-25.44） | 0104 |
| **Vascular risk factors** |  |  |  |  |
| Hypertension | 2.08 (0.60-7.15) | 0.247 | **· · ·** | **· · ·** |
| Diabetes mellitus | 0.52 (0.20-1.38) | 0.191 | **· · ·** | **· · ·** |
| Hyperlipidemia | 1.09 (0.42-2.83) | 0.867 | **· · ·** | **· · ·** |
| Coronary artery disease | 1.53 (0.37-6.26) | 0.554 | **· · ·** | **· · ·** |
| Smoking | 0.52 (0.16-1.62) | 0.256 | **· · ·** | **· · ·** |
| History of stroke | 0.88 (0.30-2.53) | 0.805 | **· · ·** | **· · ·** |
| Location of occlusion site | 0.51 (0.18-1.43) | 0.198 | **· · ·** | **· · ·** |
| i-NIHSS | 0.59 (0.45–0.78) | <0.001** | 0.53 (0.36-0.76） | <0.001** |
| IV-tPA | 0.66 (0.25–1.74) | 0.403 | **· · ·** | **· · ·** |
| Glycoprotein IIb/IIIa use  inhibitor us | 0.42 (0.15-1.17) | 0.096* | 0.67 (0.15-3.02） | 0.601 |
| General anesthesia | 0.44 (0.17-1.16) | 0.096* | 0.49 (0.12-1.99) | 0.319 |
| **Means of recanalization** |  |  |  |  |
| Stent retrieval | 0.85 (0.32-2.24) | 0.747 | **· · ·** | **· · ·** |
| Aspiration | NA | >0.999 | **· · ·** | **· · ·** |
| Balloon dilation | 0.71 (0.15-3.44) | 0.672 | **· · ·** | **· · ·** |
| Stenting | 0.21 (0.07–0.62) | 0.005** | 0.33 (0.07-1.51) | 0.152 |
| First-choice treatment (PMT vs. PBA) | 0.30 (0.10–0.93) | 0.037** | 0.10 (0.02-0.66) | 0.017** |

OR, odds ratio; aOR, adjusted odds ratio; CI, confidence interval; i-NIHSS, National Institute of Health Stroke Scale score at admission; IV-tPA, intravenous tissue-type plasminogen activator; PBA, preferred balloon angioplasty; PMT, preferred mechanical thrombectomy; * p ≤0.10; ** p≤0.05.
