## Supplementary figures and images for "Balloon Angioplasty as the First-Choice Treatment for Intracranial Atherosclerosis-Related Emergent Large Vessel Occlusion Involving the Microcatheter “First-Pass Effect”"

### Supplementary Figure 1

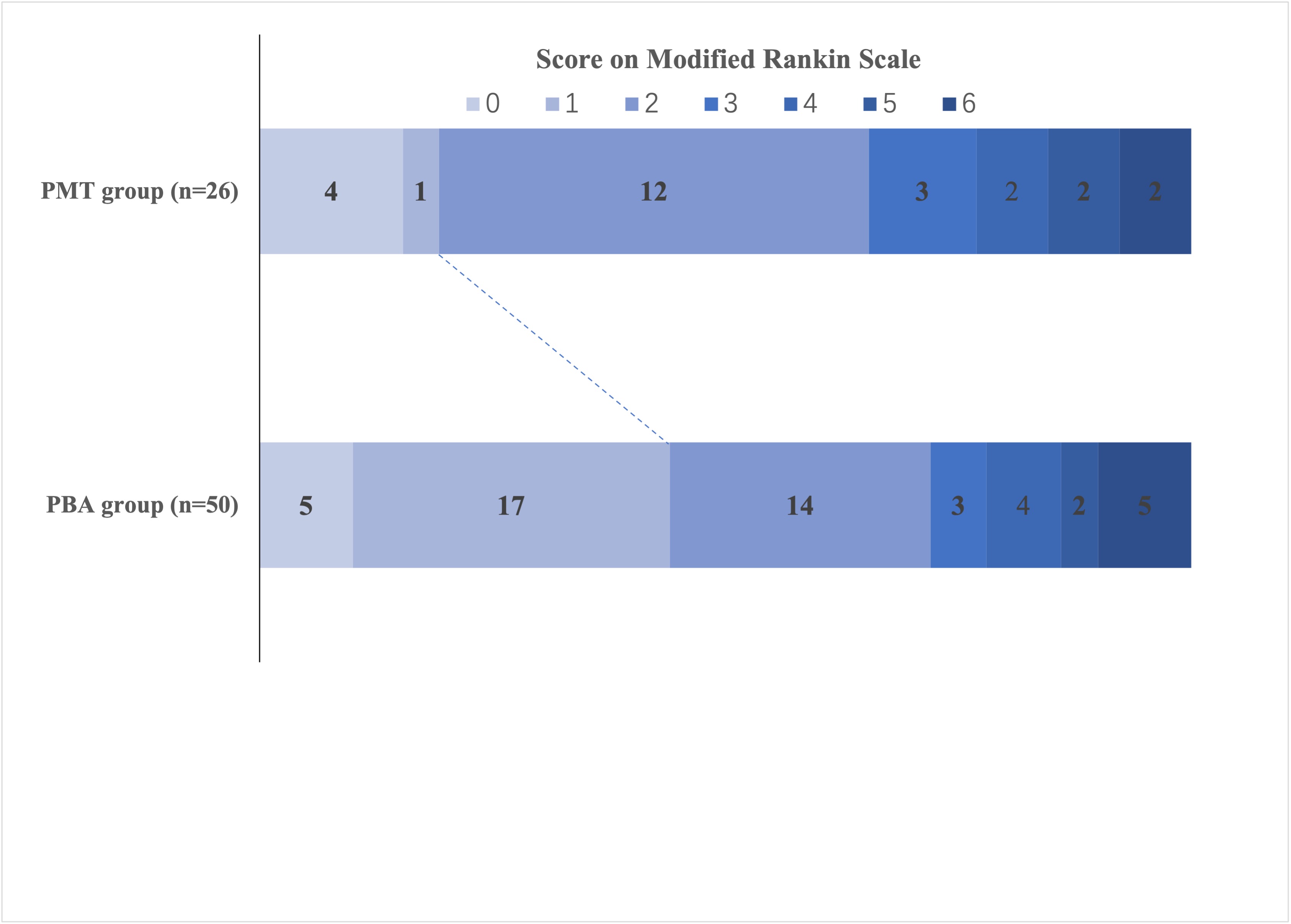
